# Accounting for cross-immunity can improve forecast accuracy during influenza epidemics

**DOI:** 10.1101/2020.07.19.20157214

**Authors:** Rahil Sachak-Patwa, Helen M Byrne, Robin N Thompson

## Abstract

Previous exposure to influenza viruses confers partial cross-immunity against future infections with related strains. However, this is not always accounted for explicitly in mathematical models used for forecasting during influenza outbreaks. We show that, if an influenza outbreak is due to a strain that is similar to one that has emerged previously, then accounting for cross-immunity explicitly can improve the accuracy of real-time forecasts. To do this, we consider two infectious disease outbreak forecasting models. In the first (the “1-group model”), all individuals are assumed to be identical and partial cross-immunity is not accounted for. In the second (the “2-group model”), individuals who have previously been infected by a related strain are assumed to be less likely to experience severe disease, and therefore recover more quickly than immunologically naive individuals. We fit both models to case notification data from Japan during the 2009 H1N1 influenza pandemic, and then generate synthetic data for a future outbreak by assuming that the 2-group model represents the epidemiology of influenza infections more accurately. We use the 1-group model (as well as the 2-group model for comparison) to generate forecasts that would be obtained in real-time as the future outbreak is ongoing, using parameter values estimated from the 2009 epidemic as informative priors, motivated by the fact that without using prior information from 2009, the forecasts are highly uncertain. In the scenario that we consider, the 1-group model only produces accurate outbreak forecasts once the peak of the epidemic has passed, even when the values of important epidemiological parameters such as the lengths of the mean incubation and infectious periods are known exactly. As a result, it is necessary to use the more epidemiologically realistic 2-group model to generate accurate forecasts. Accounting for partial cross-immunity driven by exposures in previous outbreaks explicitly is expected to improve the accuracy of epidemiological modelling forecasts during influenza outbreaks.

## 1 Introduction

Three major influenza pandemics have occurred in the 20th century, in 1918, 1957, and 1968 (Kilbourne, 2006). Each pandemic resulted in over a million deaths, with the death toll of the 1918 Spanish Flu pandemic estimated to be 50 million people (Johnson and Mueller, 2002). In 2009, a new strain of the H1N1 virus emerged, due to a reassortment of two swine viruses, triggering the first influenza pandemic of the 21st century (Trifonov et al., 2009; Christman et al., 2011). The virus is believed to have originated in Mexico in April 2009, and then spread rapidly across the globe, reaching 43 countries by May that year (Fraser et al., 2009; Trifonov et al., 2009). The case fatality rate due to the virus was lower than that of previous global pandemics in the 20th century (Kamigaki and Oshitani, 2009). However the scale of the pandemic, with estimates that 11-21% of the global population contracted the virus, significantly burdened healthcare systems (Kelly et al., 2011).

Influenza A viruses mutate over time; antigenic drift produces closely related strains, while antigenic shift causes major changes in the virus (Bouvier and Palese, 2008; Kim et al., 2018). Due to the random nature of the evolution of influenza viruses, it is not currently possible to predict when future pandemics will occur, and which strains will cause these pandemics (Neumann and Kawaoka, 2019). However, mathematical models have been used extensively for forecasting and informing public health measures when influenza outbreaks are ongoing (Ferguson et al., 2006; Hall et al., 2007; Nishiura, 2011; Ohkusa et al., 2011; Tizzoni et al., 2012; Biggerstaff et al., 2016; Thompson and Brooks-Pollock, 2019). Similarly, mathematical models are currently being used to predict the course of the ongoing COVID-19 pandemic (Ferguson et al., 2020; Kucharski et al., 2020; Prem et al., 2020; Thompson, 2020).

The most basic infectious disease outbreak models assume that individuals are epidemiologically identical (Chowell et al., 2006; Bettencourt and Ribeiro, 2008). More complex models account for differences between individuals. For example, in many studies that aim to determine optimal vaccination strategies, populations are split into low-risk and high-risk groups (Gani et al., 2005; Dushoff et al., 2007), and spatial heterogeneity can be incorporated by partitioning individuals according to their location (Longini et al., 2004; Ohkusa et al., 2009). Commonly, due to different rates of contact between individuals of different ages, as well as varying case fatality rates between age groups, age-structured models are used (Chowell et al., 2009; Medlock and Galvani, 2009; Glasser et al., 2010; Klepac et al., 2018).

Other types of heterogeneity are also likely to play an important role in the dynamics of influenza outbreaks. There is evidence that previous exposure to an influenza virus confers partial immunity to the same or similar strains, and that this protection is lifelong (Gostic et al., 2016, 2019). This partial cross-immunity may explain why there has not been a global influenza pandemic as severe as the 1918 pandemic in the last century (Thompson et al., 2019). It has been shown that a significant proportion of elderly individuals carried pre-existing immunity to the 2009 H1N1 virus (Hancock et al., 2009; Xing and Cardona, 2009; Bandaranayake et al., 2010; Hardelid et al., 2010; Gostic et al., 2019). This may be due to the similarities between the 2009 H1N1 virus and the 1918 Spanish Flu virus, as descendants of the 1918 Spanish Flu virus continued to circulate until the 1957 pandemic (Xu et al., 2010). The consequences of pre-existing immunity can be seen in the age distribution of infected individuals in Japan in the 2009 pandemic, where only a small proportion of the individuals who sought medical attention were elderly (Mizumoto et al., 2013). As well as the heterogeneity between hosts in infection risk and age mentioned previously, models in which populations are structured according to whether or not individuals carry pre-existing immunity can also be formulated (Andreasen et al., 1997; Martcheva and Pilyugin, 2006; Reluga et al., 2008; Thompson et al., 2019).

In this paper, our attention is directed towards how partial cross-immunity affects the predictability of outbreaks. We use mathematical models to investigate whether or not it is necessary to account for partially immune individuals in the population when forecasting the dynamics of future influenza epidemics. We consider two epidemiological models. In the first, partial cross-immunity is ignored (the “1-group model”). In the second, more epidemiologically realistic model (the “2-group model”), individuals with and without partial cross-immunity are accounted for explicitly.

First, we estimate values of parameters of each model (specifically, the transmission rate and the effective population size) using data from the 2009 H1N1 influenza epidemic in Japan. We then consider a synthetic future influenza outbreak of a related strain, simulated using the more epidemiologically realistic 2-group model. We explore whether or not accurate forecasts of the epidemic can be obtained in real-time. If uninformative priors are used and parameters are estimated in real-time, even the more realistic 2-group model is unable to generate accurate forecasts of the remainder of the epidemic before the peak occurs. This motivates us to incorporate information from the 2009 epidemic to set informative priors. We show that forecasts made using the 1-group model in advance or right at the start of a future epidemic are inaccurate because the model does not account for the changing number of partially immune individuals over time. We then use both information from the 2009 epidemic and data obtained as the future outbreak is ongoing to make real-time forecasts. Early in the outbreak, only the 2-group model can provide accurate forecasts of the remainder of the epidemic. For that reason, cross-immunity should be included in epidemiological forecasting models whenever an influenza outbreak is related to a strain that has previously caused a major epidemic.

## 2 Methods

### 2.1 Data

Our analysis was based on data from the 2009 H1N1 influenza epidemic in Japan comprising the estimated numbers of weekly cases seeking medical attention in that country. These data were based on data from 4800 randomly sampled sentinel hospitals, extrapolated to the total number of medical facilities in Japan (Nishiura, 2011). The data were acquired from Figure 1 of the analysis by Nishiura (2011) using the data extraction tool https://automeris.io/WebPlotDigitizer/ and the extracted data are available in Supplementary Data S1. The data represent incident cases of patients who sought medical attention and met one of the following criteria: (i) acute course of illness, (ii) fever higher than 38^°^C, (iii) cough, sputum or breathlessness (symptoms of upper respiratory infection), (iv) general fatigue, and (v) positive laboratory diagnosis.

**Figure 1:**
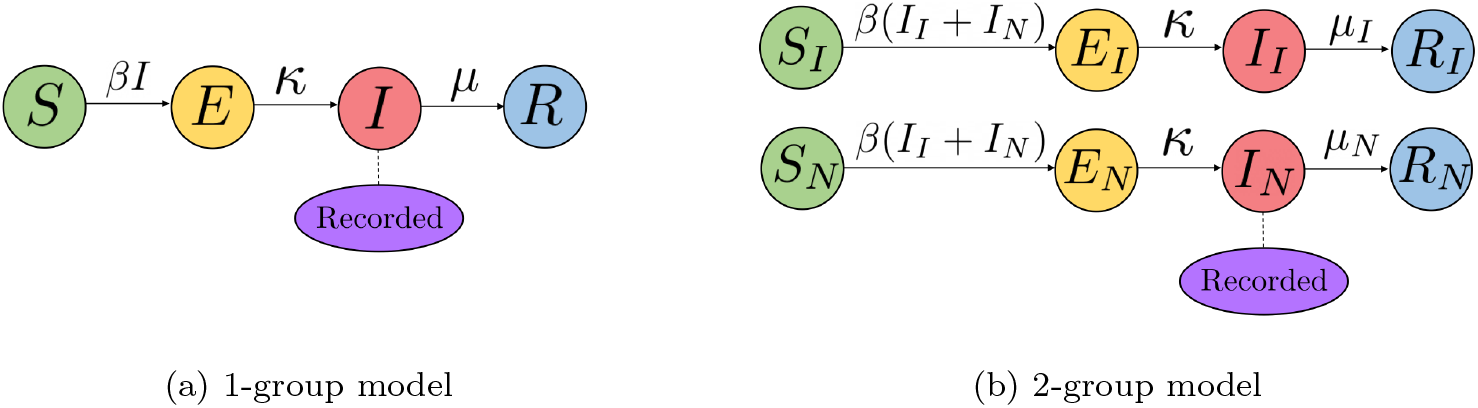
Schematics of the 1-group and 2-group models. In both cases, the population is compartmentalised into susceptible *S*, exposed (infected but not yet infectious) *E*, infected *I*, and recovered *R* classes. The 2-group model distinguishes between partially immune and immunologically naive individuals, and assumes that only infected naive individuals are recorded in case notification data (with perfect reporting). The 1-group model does not distinguish between partially immune and immunologically naive individuals, and assumes that all infected individuals are recorded in case data (again with perfect reporting).

It was estimated that 23.5% of the Japanese population was infected during the epidemic, and that 16.1% was infected and sought medical attention (Mizumoto et al., 2013). Therefore (23.5 − 16.1)/23.5 = 31.5% of infected individuals did not seek medical attention. We assume that those infected individuals who did not seek medical attention suffered mild symptoms of influenza because they were partially immune to the virus (see discussion). Hence, extrapolating to the rest of the population and assuming that the susceptibility of hosts is unaffected by partial immunity, we assume when fitting the 2-group model that 31.5% of the population were partially immune to the virus and that 68.5% were immunologically naive.

### 2.2 Models

We consider two models characterising influenza outbreaks. In the first (the 1-group model), which is the commonly used SEIR model (Anderson and May, 1991; Mills et al., 2004; Chowell et al., 2006; Chen and Liao, 2008), partial cross-immunity is neglected. In the second (the 2-group model), individuals who have been infected previously by a related strain are assumed to recover from infection more quickly than individuals who are immunologically naive. Schematics of both models are shown in Figure 1.

#### 2.2.1 1-Group Model

The 1-group SEIR model is described by the following differential equations, in which individuals are either (*S*)usceptible and available for infection, (*E*)xposed (i.e. infected but not yet infectious or symptomatic), (*I*)nfectious or, (*R*)emoved:

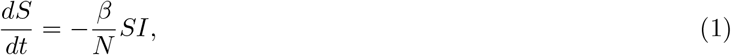

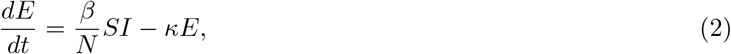

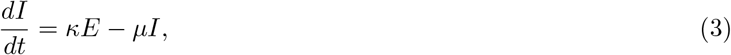

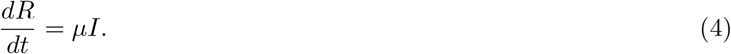

In this model, the infection rate is governed by the parameter *β*, the mean latent period is 1/*κ* weeks and the mean infectious period is 1/*µ* weeks. The basic reproduction number *R*_0_ of the 1-group model is given by

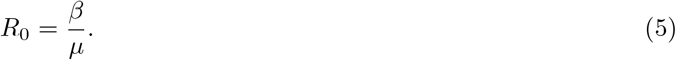

Following Cintrón-Arias et al. (2009), the number of recorded cases in week *j* (recorded at the end of that week) where *j* is the integer number of weeks since the epidemic began, is given by

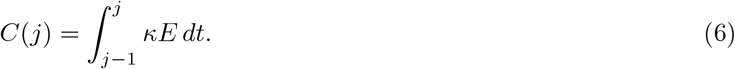

The constant value *S* + *E* + *I* + *R* = *N* represents the effective population size. Since pathogens are most likely to be transmitted locally, individuals in distant locations are not available for infection and so *N* is expected to be smaller than the true population size (Gart, 1968; Pouillot et al., 2008). In Table 1 we list the parameters that appear in equations (1)-(4) and estimates of their values for the Japanese 2009 H1N1 epidemic (see also Section 2.3 and 3.1).

**Table 1:** Descriptions of parameters of the 1-group and 2-group models and estimates of their values for the Japanese 2009 H1N1 epidemic.

| Parameter | Description | Values | Source |
| --- | --- | --- | --- |
| $\beta$ | Transmission rate | 1.644 Weeks<sup>-1</sup> | Estimated from data (Figure 3) |
| $N$ | Effective population size | $3.072 \times 10^7$ | Estimated from data (Figure 3) |
| $1/\kappa$ | Latent period | 4/7 Weeks | Tuite et al. (2009) |
| $1/\mu$ | Infectious period | 1 Week | Tuite et al. (2009) |
| $C_0$ | Initial number of recorded cases | 24073 | Mizumoto et al. (2013) |

(a): 1-group model
| Parameter | Description | Values | Source |
| --- | --- | --- | --- |
| $\beta$ | Transmission rate | 1.947 Weeks<sup>-1</sup> | Estimated from data (Figure 3) |
| $N$ | Total effective population size | $4.660 \times 10^7$ | Estimated from data (Figure 3) |
| $\nu$ | Partially immune fraction | 0.3149 | Mizumoto et al. (2013) |
| $1/\kappa$ | Latent period | 4/7 Weeks | Tuite et al. (2009) |
| $1/\mu_I$ | Infectious period (partially immune individuals) | 3/7 Weeks | Estimate based on (Fielding et al., 2013) |
| $1/\mu_N$ | Infectious period (naive individuals) | 1 Week | Tuite et al. (2009) |
| $C_0$ | Initial number of recorded cases | 24073 | Mizumoto et al. (2013) |

(b): 2-group model

#### 2.2.2 2-Group Model

The 2-group model is an extension of the standard SEIR model in which immunologically naive and partially immune individuals are distinguished between. The 2-group model is given by the following system of differential equations:

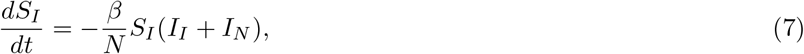

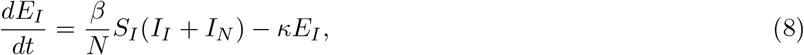

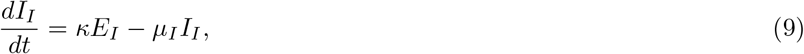

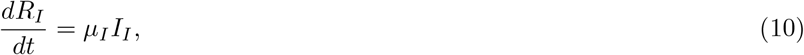

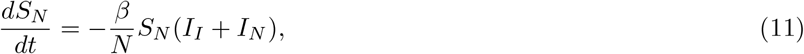

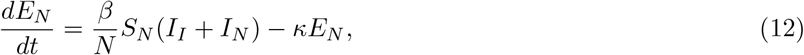

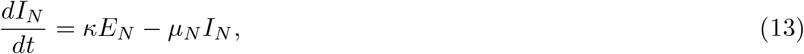

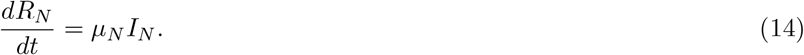

The basic reproduction number *R*_0_ of the 2-group model is given by

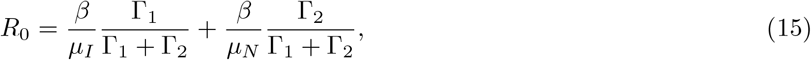

where Γ = (Γ_1_, Γ_2_) is the eigenvector corresponding to the dominant eigenvalue of the matrix

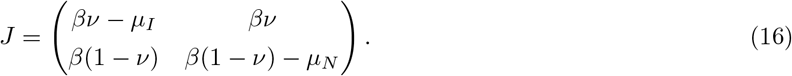

It has been shown that immune imprinting with a previous related influenza strain decreased the numbers of severe cases of H1N1, H5N1 and H7N9 influenza (Gostic et al., 2016, 2019). We assume that partially immune individuals experience less severe disease and therefore typically recover more quickly than immunologically naive individuals (i.e. 1/*µ*_*I*_ < 1/*µ*_*N*_). To isolate this effect alone on the predictability of epidemics, in this model it is assumed that partially immune and naive individuals are otherwise identical.

We assume that only cases of severe disease (i.e. infected individuals who were previously immunologically naive) report infection, so that the number of recorded cases in week *j* is given by

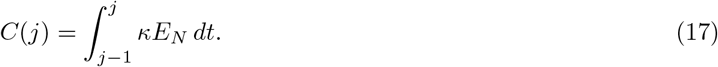

Since we assume that only immunologically naive individuals report infection, the infectious period of immunologically naive individuals in the 2-group model is assumed to be identical to the infectious period of individuals in the 1-group model (i.e. 1/*µ*_*N*_ = 1/*µ*).

Denoting the fraction of the population that is partially immune by *ν*, we have that *S*_*I*_ + *E*_*I*_ + *I*_*I*_ + *R*_*I*_ = *νN* and *S*_*N*_ + *E*_*N*_ + *I*_*N*_ + *R*_*N*_ = (1 − *ν*)*N*, where *S*_*I*_ + *E*_*I*_ + *I*_*I*_ + *R*_*I*_ + *S*_*N*_ + *E*_*N*_ + *I*_*N*_ + *R*_*N*_ = *N* is the total effective population size. In Table 1 we list the parameters that appear in equations (7)-(14) and estimates of their values for the Japanese 2009 H1N1 epidemic (again see also Section 2.3 and 3.1).

### 2.3 Parameter Estimation and Forecasting

When fitting the models to each dataset in this study, the transmission rate parameter, *β*, and effective population size, *N*, are estimated using Markov chain Monte Carlo (MCMC) with the Metropolis-Hastings algorithm (Hastings, 1970). All other parameters are assumed to be known. A likelihood function is used where it is assumed that the differences between the data and model forecasts (where the differences are due to noise not accounted for in the models) are normally distributed, and that this noise scales with the square root of the size of the data (i.e. the number of cases). We estimate this noise scaling parameter *σ* in the likelihood function (which we then fix throughout this study), by first fitting each model to data from the 2009 Japan epidemic using a least squares approach (Cintrón-Arias et al., 2009). An analysis of the residuals (the difference between the model values and data) is given in Supplementary Information Section S.1, justifying the square root noise scaling assumption. In each MCMC simulation, we perform 2 × 10^5^ sampling iterations, discarding the first 10^4^ iterations as the ‘burn-in’ period and record every 100 iterations thereafter to reduce autocorrelation. Further details are given in Supplementary Information Section S.2.

When making forecasts in real-time after *t* = *m* weeks of the epidemic has passed, we calibrate our model forecasts with the observed data for weeks 0, 1, …, *m* of the epidemic, estimating model parameters using the method just described. To generate forecasts, we re-calibrate our models to the last observed number of recorded cases at week *m*, estimating the initial conditions from the entirety of the epidemic so far, up to and including week *m* (for details, see Supplementary Information Section S.3).

### 2.4 Modelling the Size of the Partially Immune Population between Epidemics

For the 2-group model, we assume that all immunologically naive individuals infected during an earlier epidemic acquire partial immunity to related strains of the virus (and that partially immune individuals who were infected remain partially immune to related strains). We assume further that once an individual acquires partial immunity, they remain partially immune throughout their lifetime (Gostic et al., 2016, 2019). Using these assumptions, we can model the partially immune fraction of the population over the years between the end of a first epidemic and the start of a future epidemic of a related strain of influenza. We assume the next epidemic infects the same effective population as the first one (in 2009). This immune fraction of the population will decay due to births and deaths, which we now consider because the timespan between major influenza epidemics (which are often part of global pandemics) is typically many years (Kilbourne, 2006). Further details are given in Supplementary Information Section S.4. We seed the future epidemics by assuming there are 10,000 recorded cases in the first week from which we generate forecasts (three orders of magnitude smaller than the effective population sizes used in our models). When using the 2-group model to forecast future epidemics, we assume we know the partial immune fraction of the population exactly.

### 2.5 Approaches to Forecasting a Future Epidemic

In Figure 1 we have presented two mathematical models which can be used to forecast influenza epidemics. As well as determining which model to use, we shall investigate three forecasting approaches (see Figure 2 for a schematic of these approaches).

**Figure 2:**
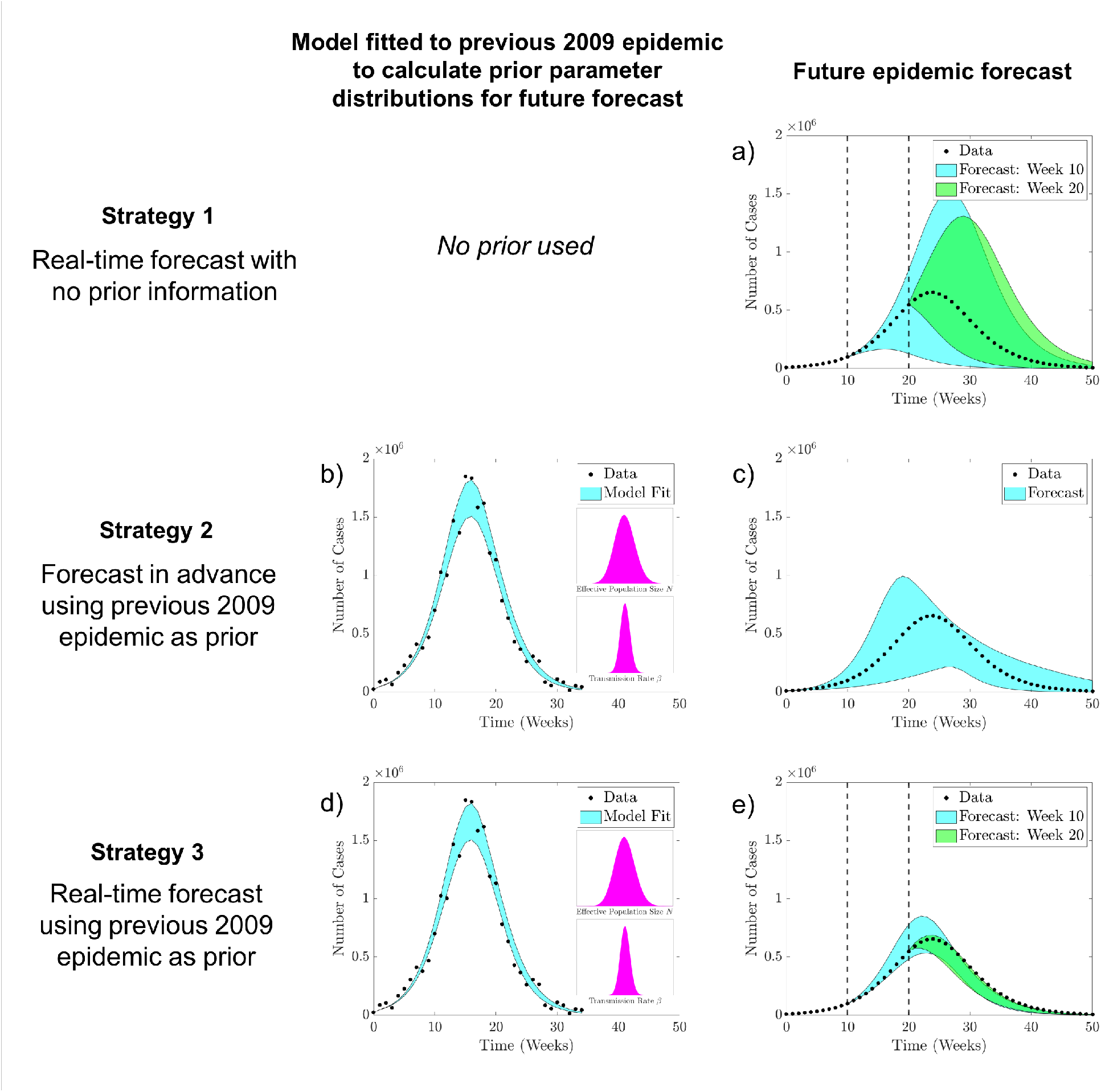
Schematic of the modelling approaches to forecast the dynamics of a future epidemic. The shaded regions indicate the 95% confidence intervals of epidemic curve trajectories and forecasts. (a) Strategy 1: Forecast in real-time, using data from the ongoing epidemic and assuming no prior information about the fitting parameters. Cyan and green forecasts calibrated using data up to week 10 and 20 of the epidemic respectively. (b)-(c) Strategy 2: Forecast in advance of a future epidemic using prior parameter distributions estimated from fitting the model to data from a previous epidemic (priors shown in panel inside (b)). (d)-(e) Strategy 3: Forecast in real-time, using data from the ongoing epidemic as well as prior parameter distributions estimated from fitting the model to data from a previous epidemic.

## 3 Results

### 3.1 Fitting Models to the 2009 H1N1 Influenza Epidemic in Japan

We fit the 1-group and 2-group models to data from the 2009 H1N1 influenza epidemic (Figure 3 (a), (b)). The shaded regions represent the 95% confidence intervals of epidemic curve trajectories based on the posterior distributions of fitting parameters *β* and *N*. Since the 1-group model does not account for infected but unrecorded individuals, the estimated parameters for the model represent a lower effective population size *N* and basic reproduction number *R*_0_ than the analogous estimates for the 2-group model (Figure 3 (c), (d)). The mean estimated values of *R*_0_ of the 1- and 2-group models are 1.644 (95%CI [1.644, 1.653]) and 1.720 (95%CI [1.704, 1.736]) respectively, comparable with the interquartile range of 1.30 to 1.70 based on fifty-seven studies of the 2009 H1N1 pandemic strain (Biggerstaff et al., 2014). The values for the estimated parameters and confidence intervals are stated in Table 2. The numbers of recorded, unrecorded, and combined total weekly cases estimated using the 2-group model are shown in Supplementary Information Section S.5.

**Table 2:** Mean values and confidence intervals of the fitting parameter posterior distributions, of the 1-group and 2-group models, fitted to data from the 2009 H1N1 influenza epidemic in Japan.

| Parameter | Mean | 95% CI |
| --- | --- | --- |
| $\beta$ | 1.644 | [1.627, 1.662] |
| $N$ | $3.072 \times 10^7$ | $[2.905, 3.246] \times 10^7$ |

(a): 1-group model
| Parameter | Mean | 95% CI |
| --- | --- | --- |
| $\beta$ | 1.947 | [1.927, 1.966] |
| $N$ | $4.660 \times 10^7$ | $[4.388, 4.938] \times 10^7$ |

(b): 2-group model

**Figure 3:**
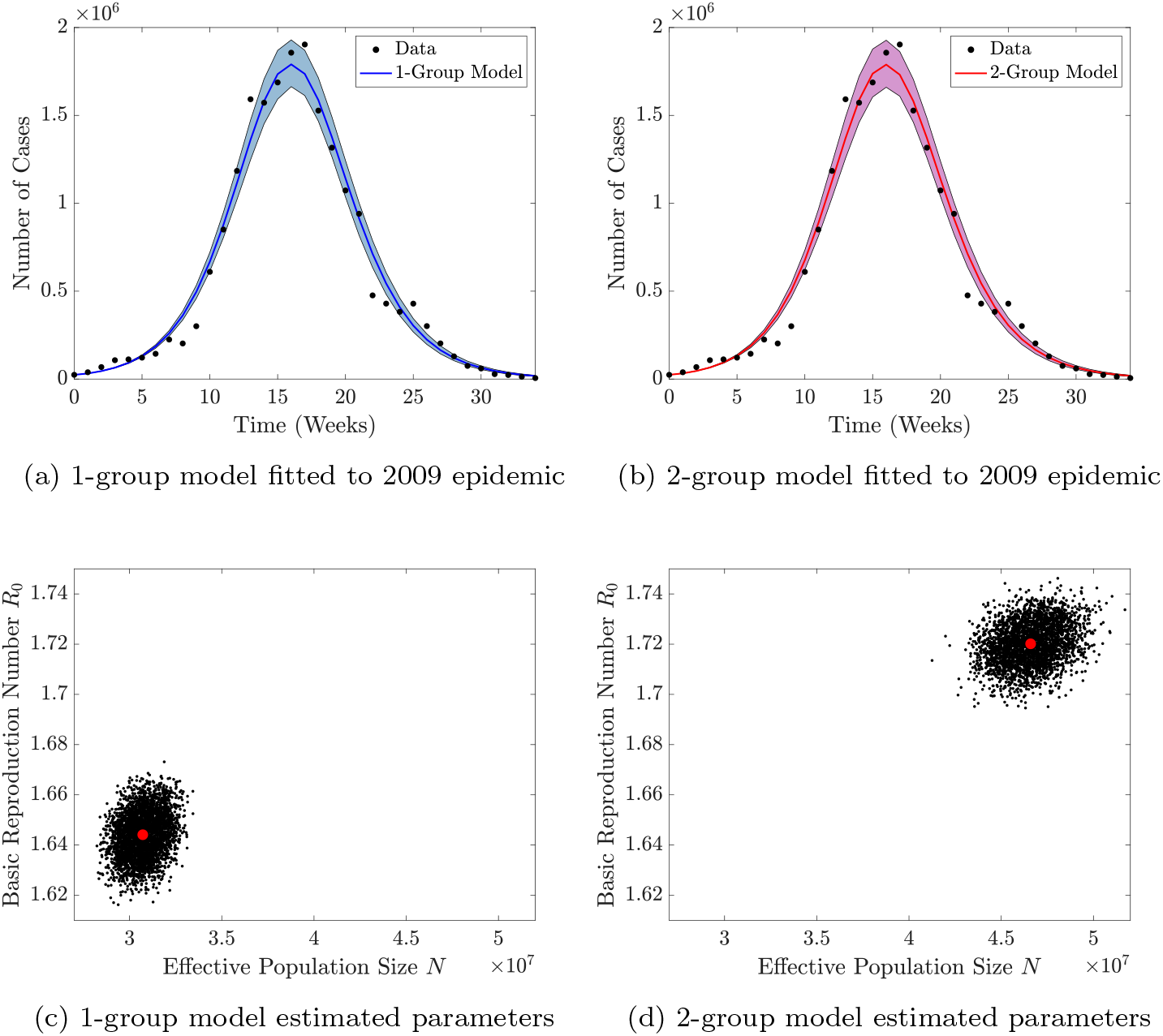
(a)-(b): The 1-group and 2-group models fitted to data of the number of new recorded cases each week from the 2009 H1N1 influenza epidemic in Japan, using the transmission rate *β* and effective population size *N* as fitting parameters. Solid lines and shaded regions indicate the mean and 95% confidence intervals of epidemic curve trajectories based on the posterior distributions of the parameters. (c)-(d): Scatter plots of posterior distributions of *R*_0_ (directly proportional to *β*) and *N*. Red dots represent the mean estimates of the parameters. Estimated parameters along with their confidence intervals are given in Table 2.

We use the mean estimated transmission rate and effective population size from the 2-group model to generate synthetic data for future epidemics. We assume that, if the simulated epidemic takes place further in future, then the immune fraction of the population is lower due to deaths of partially immune hosts and births of immunologically naive hosts (see Supplementary Information Section S.4).

### 3.2 Forecasting an Epidemic in Real-Time without Prior Information

If a major influenza epidemic were to occur, we would want to make real-time forecasts of its dynamics using live data of the number of cases each week to update predictions. We consider the case of a future epidemic occurring 25 years after the 2009 outbreak. We make forecasts using the 1- and 2-group models, at 10, 20, and 30 weeks after the first recorded cases. The results presented in Figure 4 show how the model forecasts change as we update them. The uncertainty in the forecasts of both models is large when they are made at weeks 10 or 20. By contrast, if forecasts are made at week 30, then they accurately describe the remainder of the epidemic. We remark that at week 30, the peak of the epidemic has already passed and so accurate forecasting is less practically useful. We conclude that it is difficult to forecast the dynamics of an epidemic in real-time without prior information about the transmission rate and effective population size. This result motivates us to use information from previous epidemics when forecasting the dynamics of a future one; we investigate this further below.

**Figure 4:**
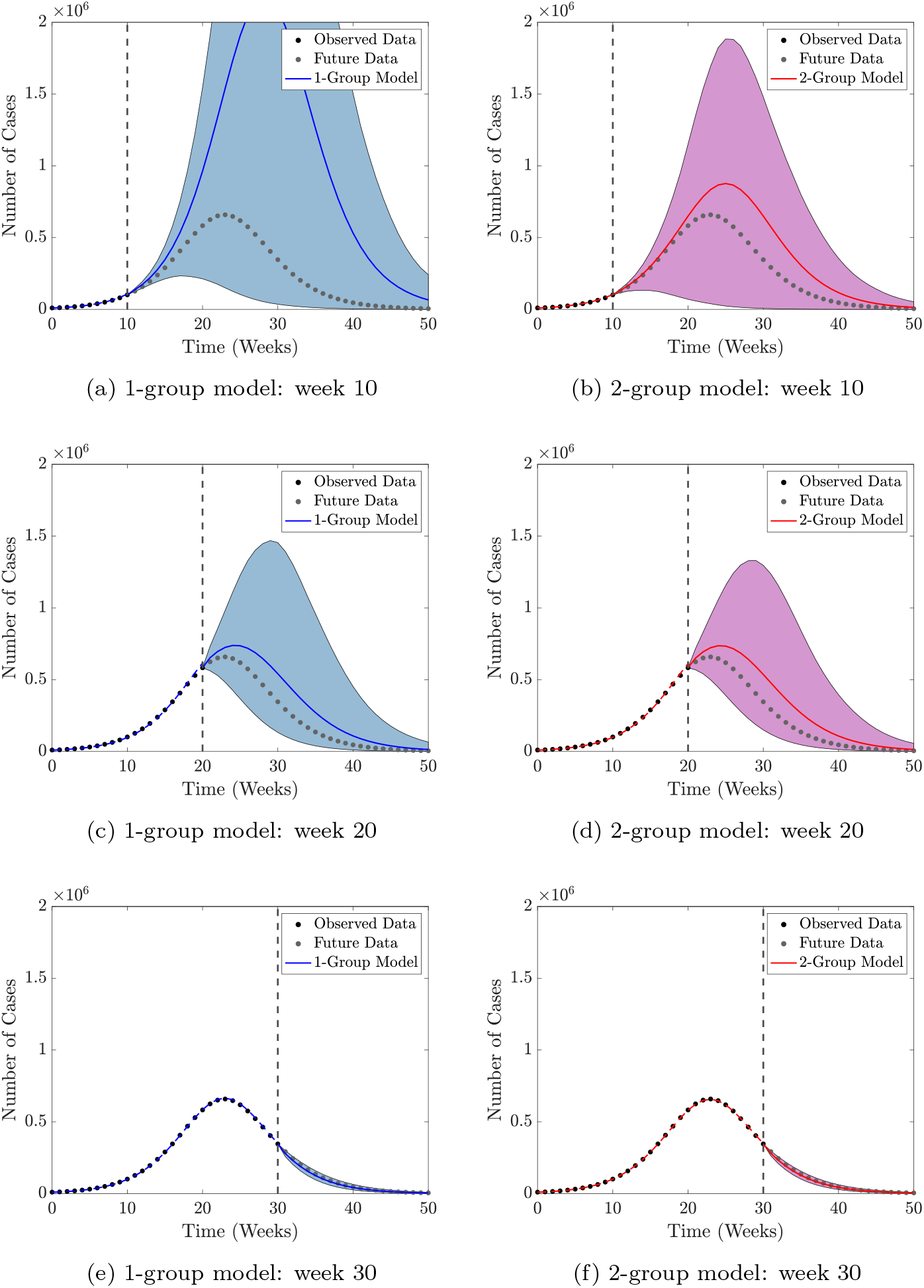
Forecast Strategy 1: real-time forecasts obtained from the 1-group and 2-group models of a future epidemic occurring 25 years after the 2009 epidemic, calibrated by fitting the model parameters *β* and *N* to data of new cases, using uninformative uniform priors. Forecasts are made at weeks 10, 20, and 30 of the epidemic, using the observed data to estimate model parameters. Vertical lines separate the calibration and forecasting periods. Dashed lines indicate the mean of the epidemic curve trajectories in the model calibration period. Solid lines and shaded regions indicate the mean and 95% confidence intervals of the forecasts, based on the posterior distributions of the parameters. The synthetic data were generated using the mean parameters of the 2-group model fitted to data from the 2009 epidemic. Uniform priors *N* ∈ [10 × 10^6^, 128 × 10^6^] and *β* ∈ [1, 3] are used.

### 3.3 Forecasting Epidemics in Advance

Using estimates of *β* and *N*, calculated by fitting the 1- and 2-group models to the 2009 epidemic, we can prescribe these parameters to follow informative gamma distributions to more accurately forecast future epidemics (values given in Supplementary Information Section S.6). We use our models to generate forecasts of the dynamics of future major influenza epidemics in Japan arising from a related strain and occurring 25, 50, or 75 years after the 2009 epidemic (with no major epidemics occurring in each intervening period). The forecasts in Figure 5 show that if the 1-group model is used where partial cross-immunity is neglected, the dynamics of a future epidemic are identical to those of the 2009 epidemic, regardless of when it occurs. By contrast, for the 2-group model, a large proportion of the population would be partially immune after the 2009 epidemic, resulting in a lower basic reproduction number. Consequently, if a related strain were to emerge 25 years later, the epidemic would be smaller than in 2009 (Figure 5 (a)-(b)). If the next major epidemic occurred 75 years later, a large proportion of the population would be immunologically naive to a related strain of the 2009 virus (because of population turnover due to births and deaths), resulting in a greater basic reproduction number. Hence, if all other factors were similar to those in 2009, the future epidemic would be larger than in 2009 (Figure 5 (e)-(f)).

**Figure 5:**
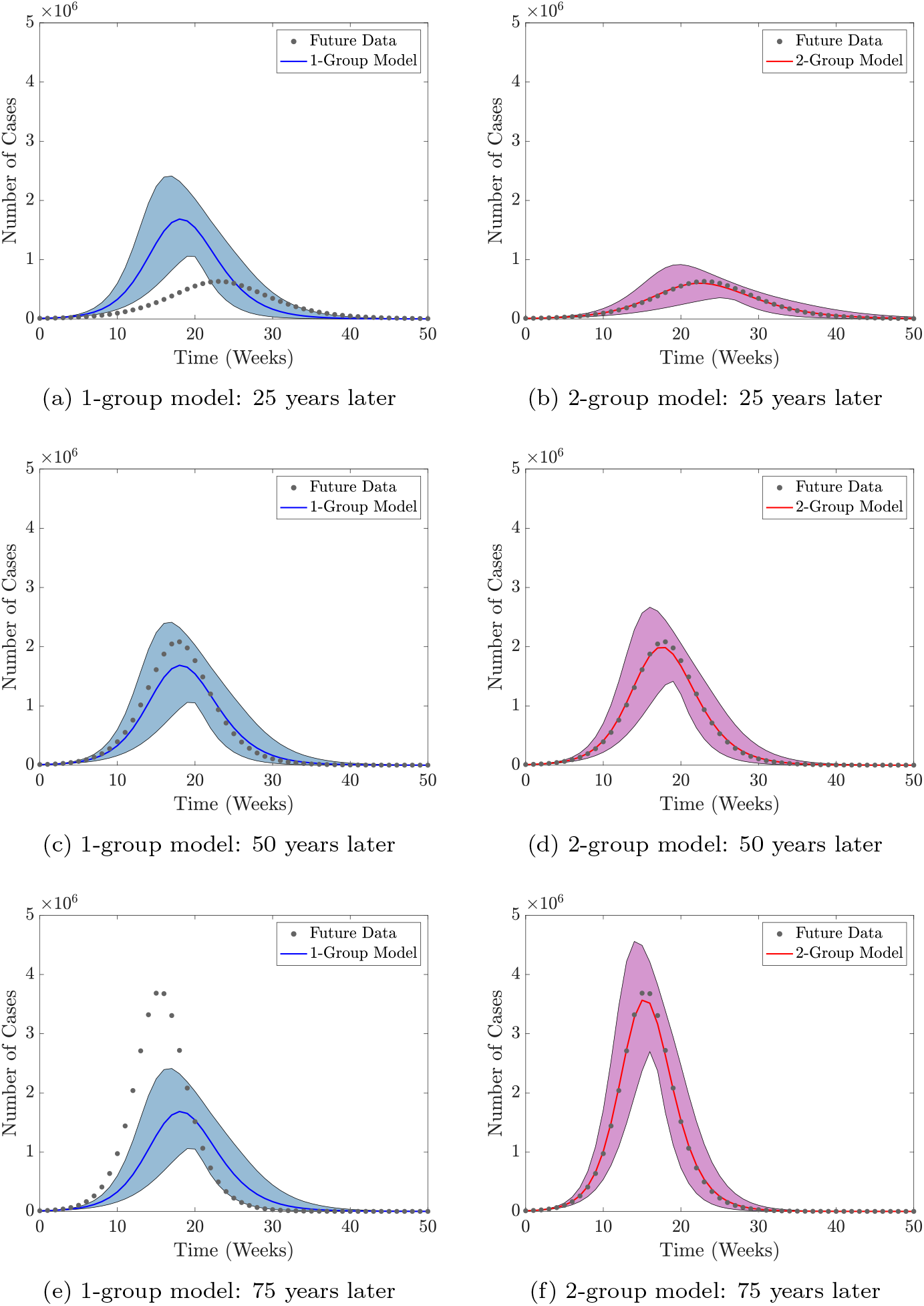
Forecasts Strategy 2: forecasts obtained from the 1-group and 2-group models of epidemics in advance, occurring 25, 50, and 75 years after the 2009 epidemic. Forecasts generated by assuming the parameters *β* and *N* follow a gamma distribution, set by fitting the models to data from the previous 2009 epidemic (details in Supplementary Information Section S.6). Solid lines and shaded regions indicate the mean and 95% confidence intervals of the forecasts. The synthetic data were generated using the mean parameters of the 2-group model fitted to data from the 2009 epidemic.

Assuming the 2-group model more accurately reflects the underlying epidemiology of the system, we conclude that the 1-group model forecasts may differ markedly from the dynamics of the next epidemic of a related strain. Forecasts using larger and smaller variances of the distributions of *β* and *N* are presented in Supplementary Information Section S.7. The partially immune fraction, basic reproduction number, total number of recorded cases, duration, and size and timing of the peak of the epidemic forecasted using the 2-group model, as the time between epidemics increases, are shown in Supplementary Information Section S.8. As seen in Figure 5, the further into the future the next epidemic of a related strain occurs then, all else being equal, the greater the total and peak number of cases and the shorter the duration of the epidemic.

### 3.4 Forecasting an Epidemic in Real-Time with Prior Information

We showed above that real-time forecasts of an influenza epidemic, without informative priors for the fitting parameters, were typically very uncertain (e.g. Figure 4 (a)-(b)). We therefore now consider using priors to inform the fitted parameter values, so that real-time forecasts are based on both historical data (from the 2009 epidemic) and live data from the ongoing outbreak. Gamma distributed priors were set for *β* and *N*, with mean values based on parameter estimates from the 2009 epidemic.

As before, we consider a scenario in which an epidemic occurs 25 years after the 2009 epidemic. In turn, predictions were made 0, 10, 20, and 30 weeks after the start of the ongoing epidemic (Figure 6 (a)–(b), (c)–(e), (f)–(h) and (i)–(k), respectively).

**Figure 6:**
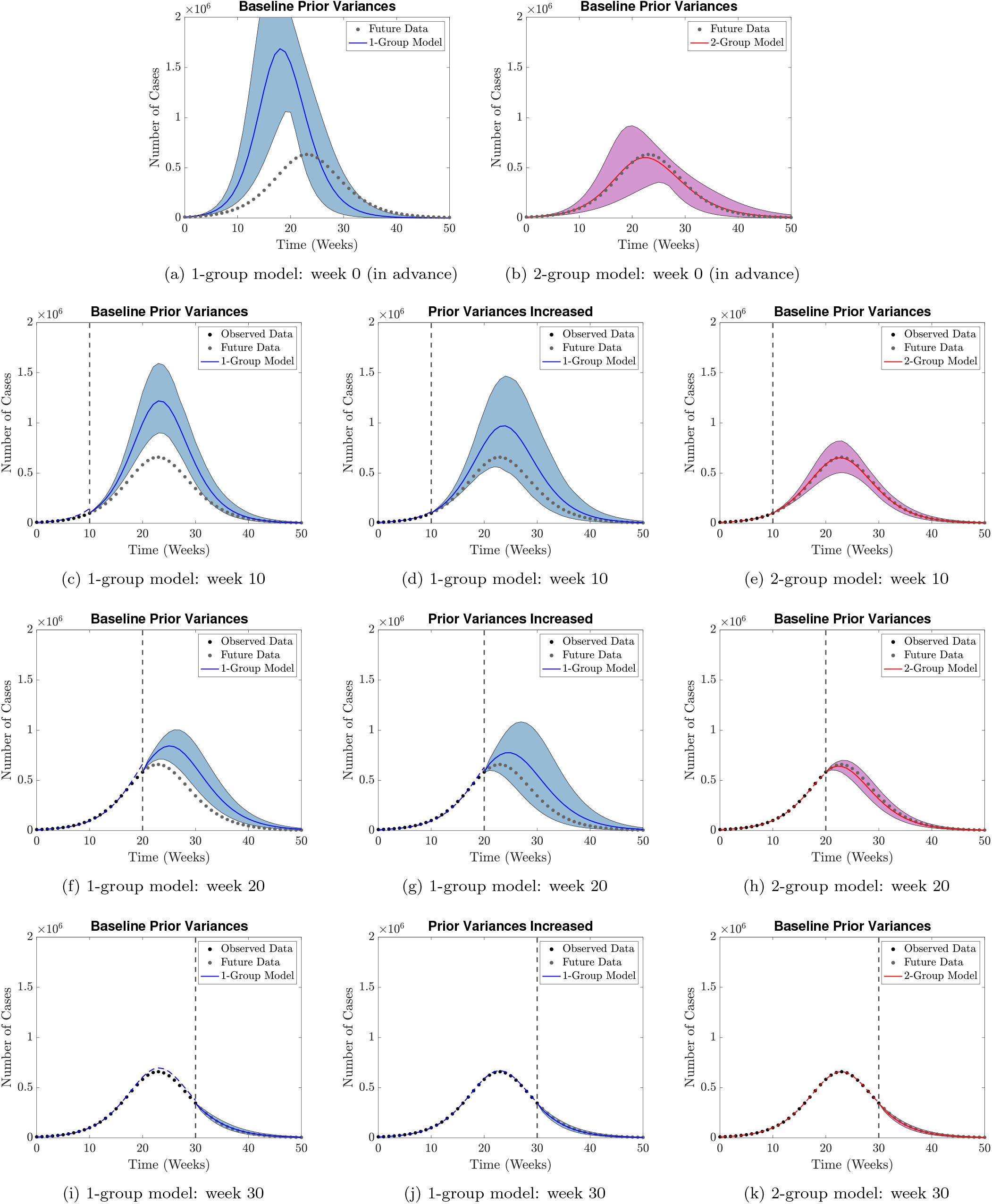
Forecast Strategy 3: forecasts obtained from the 1-group and 2-group models of a future epidemic occurring 25 years after the 2009 epidemic, in advance (at week 0) of the epidemic (a)-(b), and calibrated by fitting the models to data of new cases during the outbreak (c)-(k). Gamma distributed priors are prescribed on the fitting parameters *β* and *N*, set by fitting the models to data from the previous 2009 epidemic (details in Supplementary Information Section S.6). Forecasts are made at weeks 10, 20, and 30 of the epidemic, using the observed data to estimate model parameters. Vertical lines separate the calibration and forecasting periods. Dashed lines indicate the mean of the epidemic curve trajectories in the model calibration period. Solid lines and shaded regions indicate the mean and 95% confidence intervals of the forecasts, based on the posterior distributions of the parameters. The synthetic data were generated using the mean parameters of the 2-group model fitted to data from the 2009 epidemic.

We considered different widths of priors used to inform the epidemic forecasts (Figure 6 and figures in Supplementary Information Section S.9). Under the baseline variance considered, when the 1-group model was used, there was sometimes a discrepancy between the calibrated model trajectory and the observed epidemic data (e.g. left part of Figure 6 (f)). For that reason, we also show epidemic forecasts obtained using a wider prior (Figure 6 (d), (g) and (j)), so that the model fits the observed data more accurately. When the 1-group model is used, the forecast is either inaccurate (when a prior with low variance is used; e.g. Figure 6 (c)) or imprecise (when a prior with higher variance is used; e.g. Figure 6 (d)). In either case, our main conclusion is unchanged: predictions are improved when the more epidemiologically realistic 2-group model is used.

## 4 Discussion

Influenza epidemics, particularly pandemics, cause a significant burden on healthcare systems throughout the world (Monto, 2004). It is widely known that exposure to an influenza virus confers partial immunity to related strains (Gostic et al., 2016, 2019), and yet, partial immunity is often neglected in influenza forecasting models (Baguelin et al., 2013; Rajaram et al., 2017). In this study we have investigated whether or not it is necessary to account for partial cross-immunity when forecasting influenza epidemics and, moreover, whether data from previous epidemics can improve forecasts.

We have considered two different mathematical models which describe the spread of influenza, a 2-group model which differentiates between immunologically naive and partially immune individuals, and a 1-group model which does not. We parameterise each model using data of the estimated number of cases of infected individuals seeking medial attention per week during the 2009 H1N1 epidemic in Japan (Nishiura, 2011). When forecasting the dynamics of an epidemic of a related strain 25 years after the 2009 outbreak when there is no prior knowledge of the transmission rate and effective population size, we show that neither model predicts the dynamics of the epidemic in real-time early on during the outbreak with any certainty. Similar uncertainty was also seen when real-time forecasts were generated from an ordinary differential equation model with data from the 1918, 1957, and 1968 influenza pandemics (Hall et al., 2007).

We forecast the dynamics of future epidemics occurring at a range of time intervals after the 2009 epidemic. Immediately after the 2009 epidemic, a large fraction of the population will be partially immune to strains that are related to the 2009 H1N1 virus. This fraction reduces as the time since the 2009 epidemic increases, as individuals (a large proportion of whom carry partial immunity) die and immunologically naive individuals are born. Consequently, the size of a future epidemic of a related strain increases the further into the future at which it occurs. If we neglect partial cross-immunity by using the 1-group model, we predict the same outbreak dynamics as in 2009, regardless of when it occurs. This is because changing numbers of partially immune individuals are not accounted for. This would either result in an overestimation of the size of a future epidemic, if the partially immune fraction was greater than the baseline partial immunity in 2009 (i.e. if the future outbreak occurred soon after 2009), or an underestimation, if it was less (i.e. if the future outbreak occurred long after 2009).

Finally, we considered incorporating knowledge of parameters from the 2009 epidemic, combined with using outbreak data to make real-time forecasts during a future epidemic. When the 1-group model was used to make forecasts, we found that wide priors had to be used to calibrate the model trajectories to the data, especially early on. Although these priors, based on the 2009 epidemic, improved the forecasts compared to when no prior information was used, the forecasts were still uncertain when made before the peak of the epidemic. By contrast, the 2-group was able to make more accurate forecasts as it accounted for the changes in the number of partially immune individuals and, hence, its priors accurately reflected the values of the true underlying parameters of the system. We conclude that to forecast the long-term dynamics of major influenza epidemics, a model that accounts for partial cross-immunity should be used, and data from related previous epidemics taken into account. We note that there may be instances in which the underlying epidemiology of an outbreak is not well known, or where a complex model cannot be parameterised due to poor surveillance and or lack of data (Gibbons et al., 2014). In such cases, using a simple model akin to the 1-group model used in this study may be the only possible option for making predictions. Although forecasts made by a 1-group model may lack precision, they could still be useful to policy makers during an epidemic. For example, the 1-group model was able to accurately predict the duration of the epidemic when using data from the first 10 weeks of the outbreak, even if it could not predict well the trajectory of the entire epidemic. Such models may also be useful for short-term forecasts (Funk et al., 2019). By constantly updating model predictions using new data, and adjusting parameter assumptions when model calibration trajectories deviate from the data, simple models which do not describe the underlying epidemiology of an epidemic fully may still be informative.

We investigated whether or not it is necessary to include a particular source of heterogeneity (i.e. partial immunity) when forecasting influenza epidemics. In a similar manner, we could extend our models to question whether other sources of heterogeneity should be accounted for. For instance, age-structure could be incorporated (Nishiura et al., 2010). We could also include spatial heterogeneity, by partitioning the population into distinct geographical regions (Ohkusa et al., 2009). A significant challenge with this extension would be identifying the transmission rates between different regions.

In the 2-group model considered here, we assumed, as in Reichert et al. (2012), that immunoprotection does not prevent infection, only its consequences. Different types of partial cross-immunity to related strains of a virus can be considered. For example, in Thompson et al. (2019) it was assumed that cross-immunity decreases susceptibility, that is, the probability that an individual becomes infected, an assumption also used in influenza transmission models (Hill et al., 2019). Future work could consider whether or not the results of this study hold when different assumptions are made about the precise effects of cross-immunity. In the 2-group model, in order to study the effects of cross-immunity in as simple a setting as possible, we assumed that infected individuals who did not seek medical attention were already partially immune to the virus. However, a range of factors (including age and behaviour) are likely to affect the probability that an individual seeks medical attention. This could be built into the underlying modelling framework considered here, although additional data would be needed to parameterise the resulting model. Other additions to the 1- and 2-group models could also be considered. For example, the wide range of interventions that are implemented during outbreaks could be included in the models explicitly (Gani et al., 2005; Longini et al., 2005; Backer et al., 2019).

Nonetheless, our approach has demonstrated the principle that including partial cross-immunity in forecasting models during influenza epidemics can lead to more accurate forecasts. Cross-immunity due to previous infection has been shown to play a major role in the dynamics of influenza epidemics, with clear evidence emerging from the 1918 Spanish Flu pandemic (Taubenberger and Morens, 2006) and the 2009 H1N1 pandemic (Hancock et al., 2009). We expect cross-immunity to remain important in future influenza epidemics. Consideration of partial cross-immunity by epidemiological modellers is therefore of obvious public health importance.

## Data Availability

Data available from the ResearchGate link.

https://www.researchgate.net/publication/343058991_Supplemtary_Data_S1pdf

## Author’s Contributions

All authors conceived the study. RS-P conducted the analysis. RNT and HMB supervised the research. All authors wrote and revised the manuscript. All authors approved the final version of the manuscript.

## Acknowledgements

Thanks to Erin Lafferty and Claude Schmit (Biosensors Beyond Borders Ltd.), and the members of the Wolfson Centre for Mathematical Biology (University of Oxford) for helpful discussions about the work.

## Supplementary Information

### S.1 Residual Analysis of Models Fitted to the 2009 Epidemic

There may be situations in which the assumptions of a model underlying the parameter estimation framework are violated, for instance, how the amplitude of the noise in the data scales with the number of cases. These incorrect assumptions may not be immediately apparent when observing the fitted models and data. However, to look for any systematic deviations between the fitted models and data, we can carry out a residual analysis, plotting the residuals (the differences between the model values and observed data) against time and the model values (Cintrón-Arias et al., 2009).

In Figure S1 we plot the residuals, as calculated by (S6) of the 1-group and 2-group models with least squares estimated parameters (see Supplementary Information Section S.2), fitted to data from the 2009 H1N1 influenza epidemic in Japan, against the model values (number of cases) and time. There are no trends or any discernible patterns in any panel in Figure S1, which suggests that it is reasonable to assume that the noise in the observed data of the number of cases scales with the square root of the size of the data.

**Figure S1:**
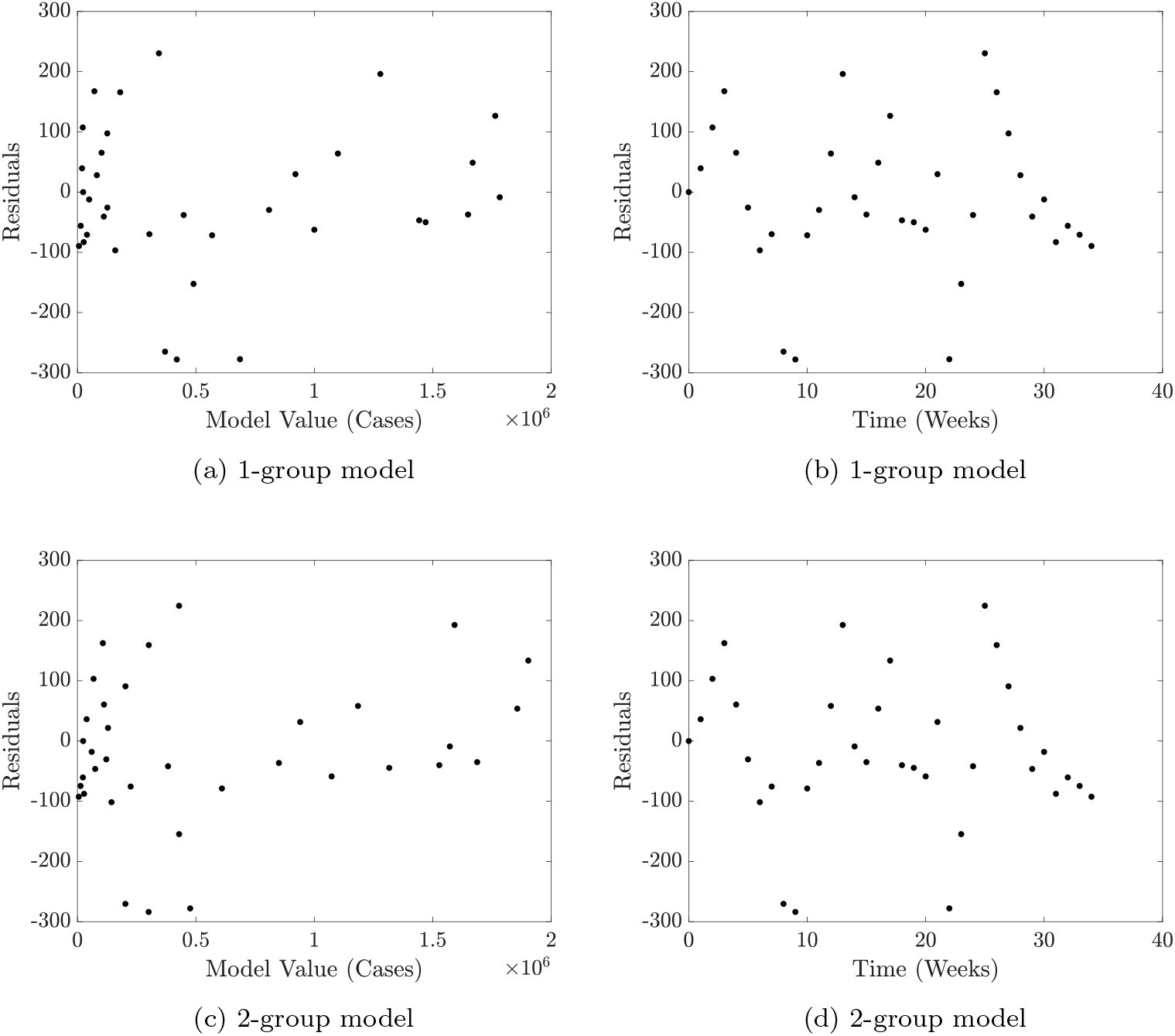
Residuals of the 1-group and 2-group models fitted to to data from the 2009 H1N1 influenza epidemic in Japan, with least squares estimated parameters, calculated using (S6), plotted against the model value and time.

### S.2 Parameter Estimation

We use Markov chain Monte Carlo (MCMC) with the Metropolis-Hastings algorithm to estimate model parameters (Hastings, 1970). It is assumed that the noise in the observed data scales with the square root of the size of the data (i.e. the number of cases), hence we use the likelihood function

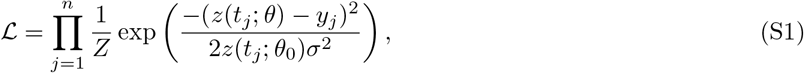

where *z*(*t*_*j*_, *θ*_0_) are the number of cases given by our model with the parameters *θ, y*_*j*_ are the observed data, and *Z* is a normalisation constant we need not consider.

To determine *σ*, we first fit the 1- and 2-group models to data from the 2009 epidemic in Japan using a least squares procedure, again assuming that the observed data scales with the square root of the size of the data, and that the observed data deviates from the model forecasts due to noise which the model does not account for, such that

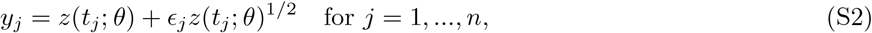

where *ϵ*_*j*_ are assumed to be independent and identically distributed random variables with zero mean *E*[*ϵ*_*j*_] = 0 and finite variance var[*ϵ*_*j*_] = *σ*^2^. We define the cost function

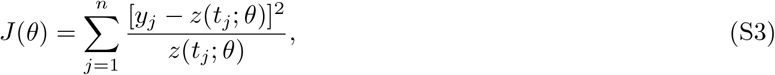

The least squares parameter estimates are given by

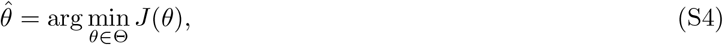

where Θ is the feasible set of parameter values. We solve the optimisation problem (S4) using MATLAB’s `fminsearch` function.

Following Cintrón-Arias et al. (2009), we estimate the noise scaling parameter *σ*^2^ by

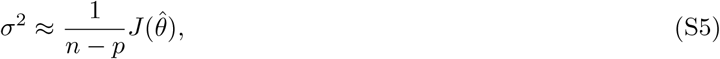

where *p* is the number of parameters estimated from the data, and define the residuals by the ratio

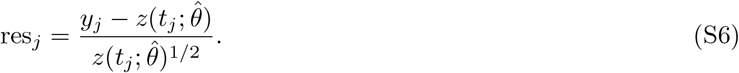

When fitting the models to data from the 2009 epidemic in Japan, we estimate *σ* using (S5), calculating *σ* = 123.5 for the 1-group model and *σ* = 123.9 for the 2-group model respectively. Hence, we use the value *σ* = 124 in the likelihood function (S1) throughout this study. A residual analysis of both models with the least squares estimated parameters is given in Supplementary Information Section S.1, justifying the assumption the noise in the observed data scales with square root size of the data.

### S.3 Estimation of Initial Conditions

The observable data we consider is the number of cases that have been recorded each week, *C*(*j*). Given that we observe *C*(*j*) for *j* = 0, 1, …, *m*, we wish to estimate the number of individuals in each compartment of our models at week *j* as initial conditions to make forward forecasts.

When estimating initial conditions from case data from multiple previous week (i.e. when we are part way through an epidemic), it is necessary to take data from every week into account, as for instance, an individual who was reported as being infected many weeks ago may not have recovered yet. As we estimate initial conditions from the observed data rather than model trajectories, we make the assumption that the cases are generated uniformly in time (e.g. if there were seven cases reported in one week, we assume that there was one new case each day).

Assuming an exponentially distributed infectious period as in our models, and first considering the 1-group model, if *C*(*j*) cases were reported each week for *j* = 0, 1, …, *m*, the number of infected individuals *I* at week *m* is given by

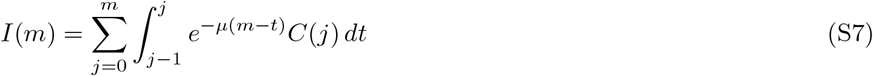

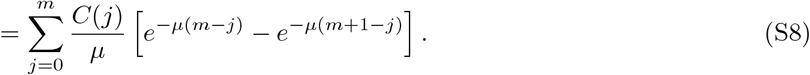

Restating (6), we have

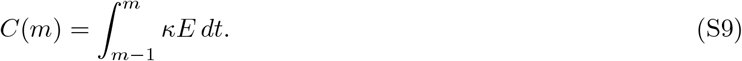

Hence, by assuming a constant *E* over the past week, we can make the estimation

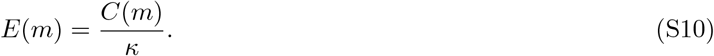

The number of newly recovered individuals each week is given by the difference between the number of new cases and the number of individuals relating to those cases who are still infected. Hence, we estimate the number of recovered individuals *R* at week *m* by

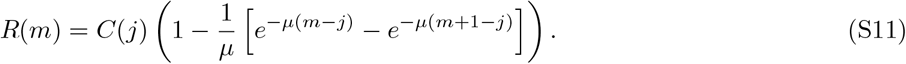

As we have assumed a total fixed effective population size *N*, the number of susceptibles at week *m* is given by

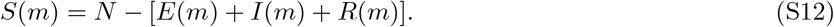

We now consider the 2-group model. As it is assumed that only cases of naive individuals becoming infected are reported, we estimate the number of infected naive individuals at week *m* to be

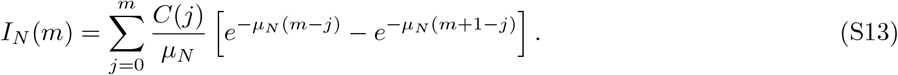

As partially immune and naive individuals are equally likely to be infected, we estimate the number of new (unrecorded) cases of infectious partially immune individuals each week to be 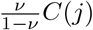. Hence, we estimate that

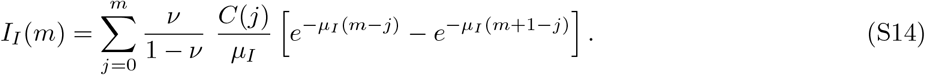

The number of exposed, recovered, and susceptible individuals at week *m* in each group are estimated by

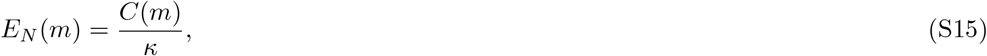

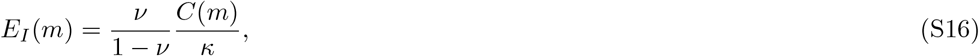

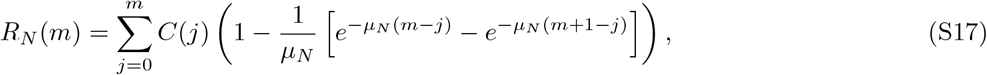

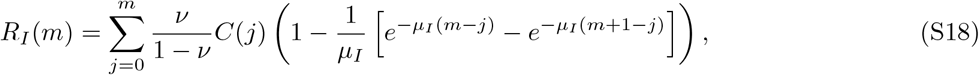

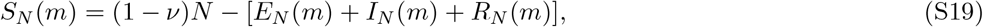

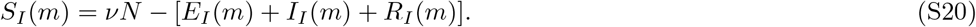

### S.4 Modelling the Partially Immune Fraction of the Population

We assume that all infected naive individuals in the 2009 H1N1 influenza epidemic in Japan acquired partial immunity to the virus and related strains. Hence, we can compute the partially immune fraction of the population immediately after the epidemic, *ν*^∗^, by

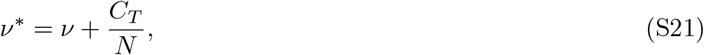

where *ν* is the immune fraction before the epidemic, *C*_*T*_ is the number of total cases recorded in the data, and *N* is the total effective population size. Hence, from the 2-group model fitted to data from the 2009 epidemic in Japan as shown in Figure 3, we estimate the partially immune fraction of the population immediately after the epidemic to be *ν*^∗^ = 0.7443, using the mean estimated total effective population size, *N* = 4.660 × 10^7^.

To determine the partially immune fraction when a second epidemic in the future begins, we shall compute the fraction as a function of the time since the end of the first epidemic. We shall assume that once an individual acquires partial immunity, they remain partially immune for their lifetime.

Using population data from Japan in 2009 accessed from National Insititute of Population and Social Secrity Research (2020), which we display in Figure S2, we define the number of individuals aged *a* by *b*(*a*), and the number of the number of deaths of individuals aged *a* by *d*(*a*). Hence, the probability, *P* (*a, n*), that an individual of age *a* survives for *n* more years is given by

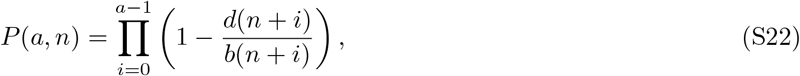

where *d*(*a*)/*b*(*a*) is the probability than an individual aged *a* dies within one year.

The fraction of individuals who have survived for *n* years is then given by

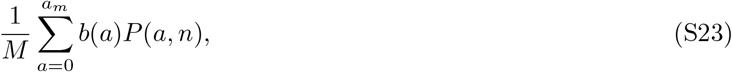

Where 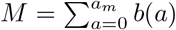, and *a*_*m*_ is the assumed maximum age of an individual, which we take to be 110. Hence assuming a constant population size and that all individuals are born immunologically naive, we can calculate the immune fraction *ν, n* years after the first epidemic to be

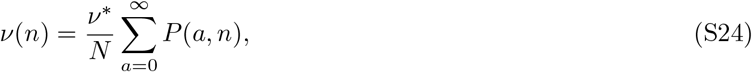

where *ν*^∗^ is the immune fraction immediately after the first epidemic. The result is shown in Figure S6 (a) (see Supplementary Information Section S.8). For an epidemic occurring 25 years after 2009 as we consider in this study, the partially immune fraction of the population at the start of the epidemic is *ν* = 0.5129.

**Figure S2:**
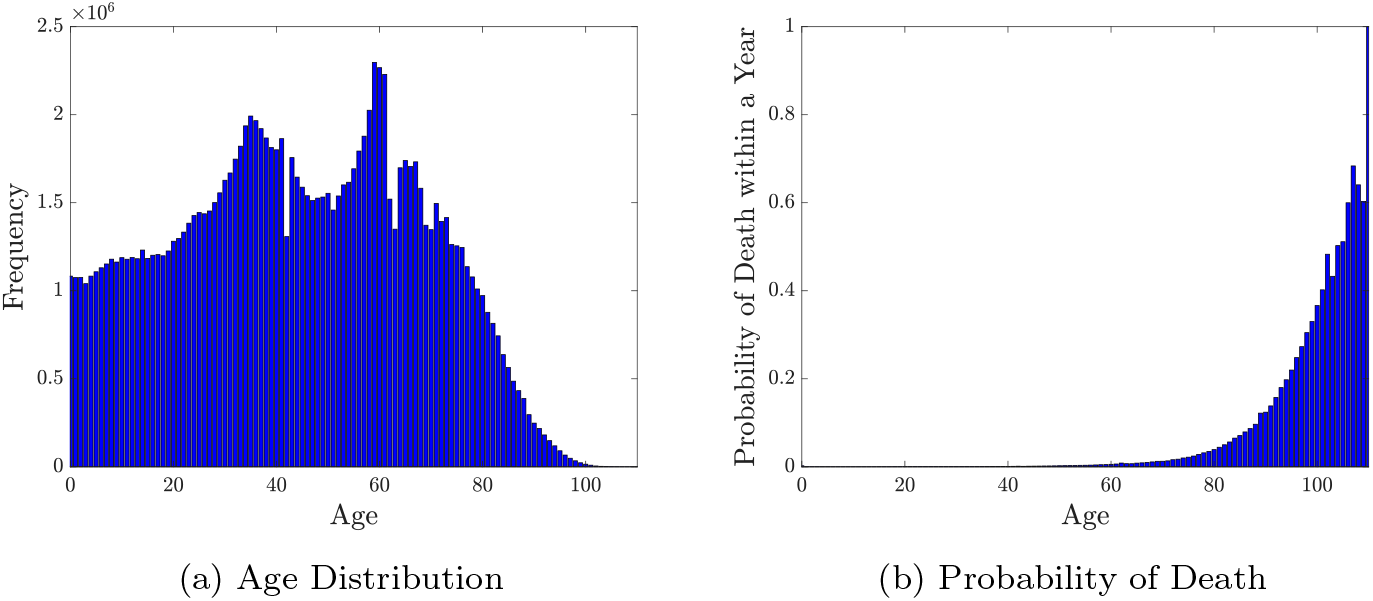
(a): The age distribution of the population of Japan in 2009. (b): The age distribution of the probability that an individual in Japan dies within on year, based on mortality and population data from 2009. Data accessed from National Insititute of Population and Social Secrity Research (2020)

### .S 2-Group Model Fitted to the 2009 Epidemic: Total Recorded and Unrecorded Cases

We assume that all cases of immunologically naive infected individuals are recorded in the data, and that no cases relating to partially immune individuals are recorded, due to the assumption that naive individuals suffer more severe symptoms and are therefore more likely to seek medical attention. In Figure S3 we display the number of (recorded) infected naive individuals, the number of (unrecorded) infected partially immune individuals, and their combined total, estimated by the 2-group model fitted to the data from the 2009 epidemic in Japan.

**Figure S3:**
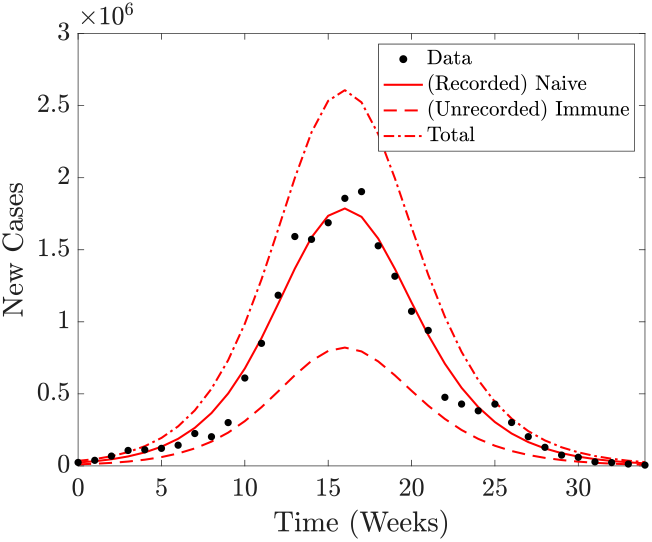
The weekly number of recorded, unrecorded, and total (recorded and unrecorded) cases as estimated by the 2-group model with mean estimated parameters, fitted to data from the 2009 Japanese influenza epidemic.

### S.6 Prior Fitting Parameter Distributions

We prescribe prior gamma parameter distributions on *β* and *N* using estimated values from the 2009 epidemic. Different variances of these distributions may be considered when making forecasts, values of which are given in Table S1.

**Table S1:** Mean and variance of prior gamma distributions of the transmission rate *β* and effective population size *N* used to make forecasts of future epidemics using the 1-group and 2-group models, set by fitting models to data from the 2009 epidemic.

| Figure | Parameter | Mean | Variance | Relative Variance |
| --- | --- | --- | --- | --- |
| 5, 6 (a), 6(c), 6(f), 6(i) | $\beta$ | 1.644 | $5 \times 10^{-3}$ | 1 |
| 5, 6 (a), 6(c), 6(f), 6(i) | $N$ | $3.063 \times 10^7$ | $1 \times 10^{13}$ | 1 |
| 6 (d), 6 (g), 6 (j), S4, S7 | $\beta$ | 1.644 | $2.5 \times 10^{-2}$ | 5 |
| 6 (d), 6 (g), 6 (j), S4, S7 | $N$ | $3.063 \times 10^7$ | $5 \times 10^{13}$ | 5 |
| S5, S8 | $\beta$ | 1.644 | $1 \times 10^{-3}$ | 0.2 |
| S5, S8 | $N$ | $3.063 \times 10^7$ | $2 \times 10^{12}$ | 0.2 |

(a): 1-group model
| Figure | Parameter | Mean | Variance | Relative Variance |
| --- | --- | --- | --- | --- |
| 5, 6 | $\beta$ | 1.947 | $5 \times 10^{-3}$ | 1 |
| 5, 6 | $N$ | $4.649 \times 10^7$ | $1 \times 10^{13}$ | 1 |
| S4, S7 | $\beta$ | 1.947 | $2.5 \times 10^{-2}$ | 5 |
| S4, S7 | $N$ | $4.649 \times 10^7$ | $5 \times 10^{13}$ | 5 |
| S5, S8 | $\beta$ | 1.947 | $1 \times 10^{-3}$ | 0.2 |
| S5, S8 | $N$ | $4.649 \times 10^7$ | $2 \times 10^{12}$ | 0.2 |

(b): 2-group model

### S.7 Forecasting Epidemics in Advance using Different Prior Parameter Distributions

Different variances of the prior parameter distributions of *β* and *N* may be considered my modellers. We show forecasts of future epidemics made in advance of future outbreaks occurring 25, 50, and 75 years after the 2009 epidemic, if variances five times larger (Figure S4) or five times smaller (Figure S5) of both *β* and *N* are used, compared to forecasts made in Figure 5.

**Figure S4:**
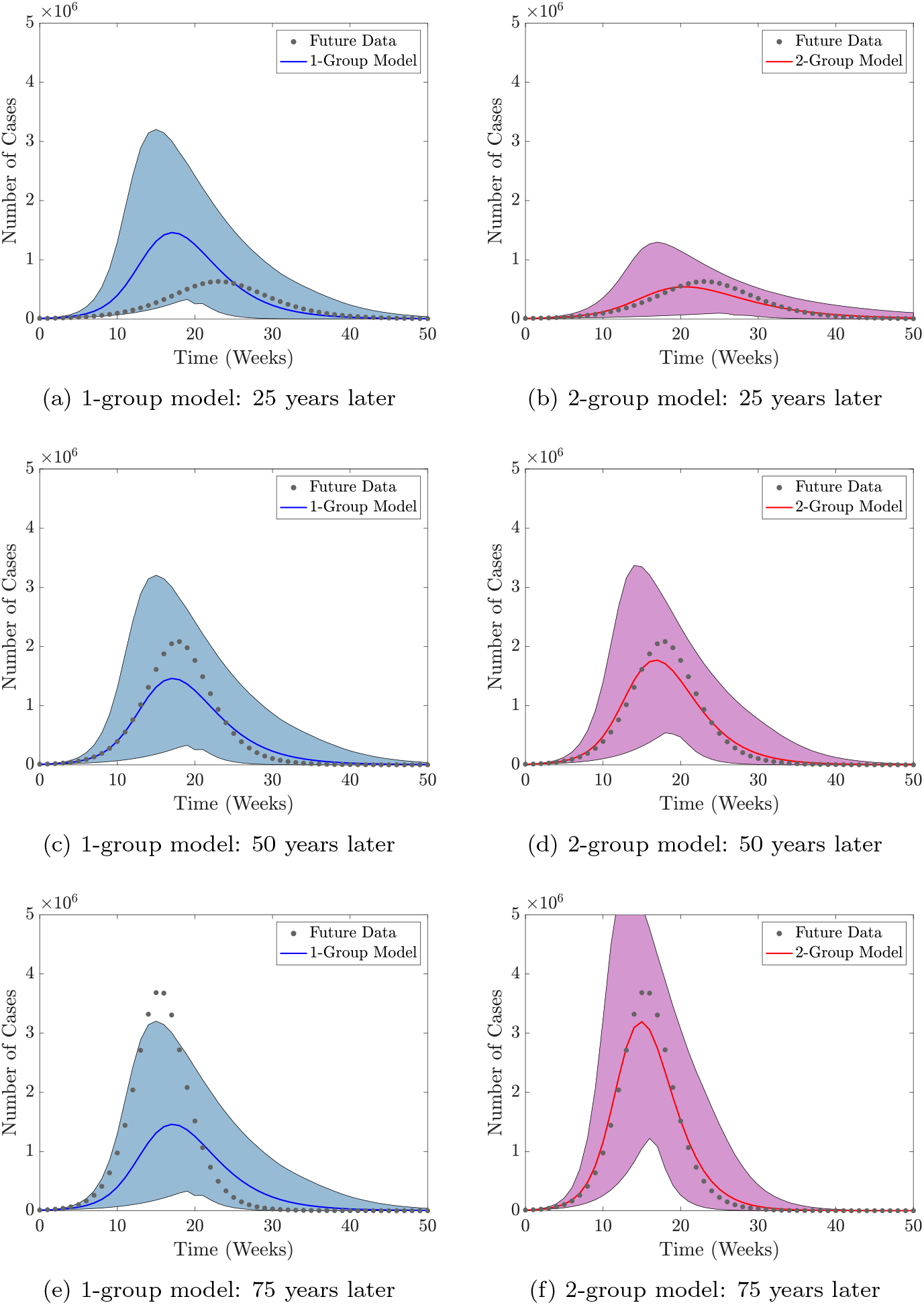
Forecasts obtained from the 1-group and 2-group models of epidemics occurring 25, 50, and 75 years after the 2009 epidemic as in Figure 5, but with variances of the gamma distributions of *β* and *N* five times larger (details in Supplementary Information Section S.6).

**Figure S5:**
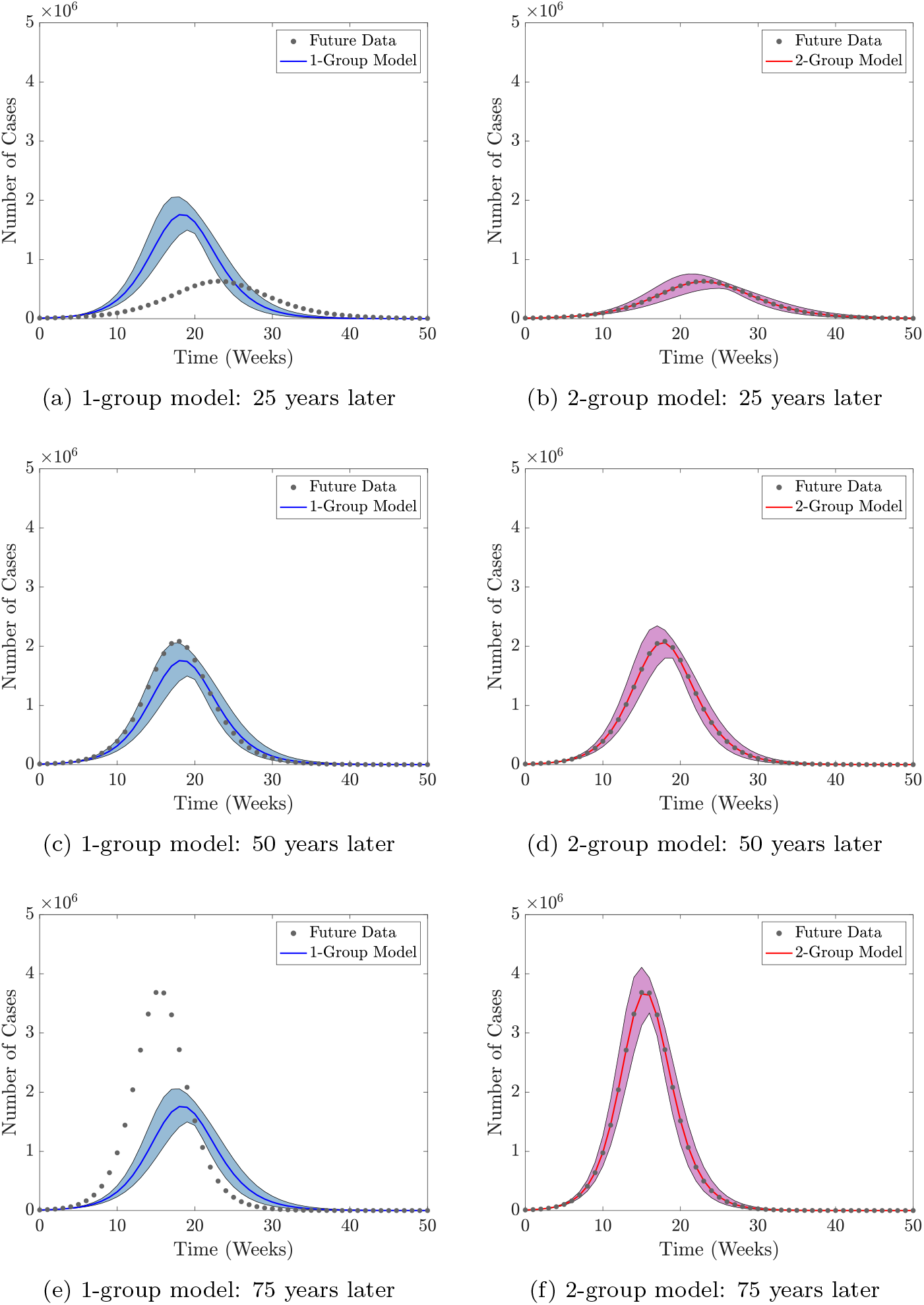
Forecasts obtained from the 1-group and 2-group models of epidemics occurring 25, 50, and 75 years after the 2009 epidemic as in Figure 5, but with variances of the gamma distributions of *β* and *N* five times smaller (details in Supplementary Information S.6).

### S.8 Outbreak Statistics of Future Epidemics

In Figure S6 we show how the partially immune fraction, basic reproduction numbers, and various quantities of interest relating to the dynamics of a future epidemic (estimated using both the 1-group and 2-group models) vary dependent on when the future epidemic occurs. The partially immune fraction decreases approximately linearly before tapering to zero as time increases (Figure S6 (a)). This results in an increase in the basic reproduction number, *R*_0_, of the 2-group model (Figure S6 (b)).

We explore various quantities of interest which would inform policy makers on how best to control the epidemic: (i) the total number of recorded cases of individuals seeking medical attention, (ii) the duration of the epidemic, (iii): the maximum weekly number of recorded cases, and (iv): the time at which the maximum weekly number of recorded cases occurs. Under the assumptions of the 1-group model, these are all forecasted to be fixed irrespective of when the next epidemic begins, and identical to that of the 2009 epidemic. However, under the assumptions of the 2-group model, these quantities all vary, which we now describe, as well as motivations for determining them.

The total number of recorded cases of individuals seeking medical attention reflects the number of total medical resources (e.g. antivirals, hospital beds, staff) required to treat individuals (Figure S6 (c)). This increases in the 2-group model the further into the future the next epidemic begins, due to the decreasing partially immune population fraction. It would be useful for public health officials and medical facilities to know the expected duration of the epidemic so they can plan and allocate resources accordingly. Here we define the duration of the epidemic to be the time until the number of recorded cases reaches fewer than 1000 per week (orders of magnitude smaller than the peak of the epidemic). The duration of the next epidemic would decrease the further into the future in occurred, and is inversely correlated to the number of cases (Figure S6 (d)). This is because if there was a greater proportion of immunologically naive individuals in the population, the basic reproduction number would be larger and hence the virus would spread more quickly leading to a shorter epidemic duration.

The maximum weekly number of recorded cases (i.e. the peak of the epidemic) and the time at which this peak occurs are important to public health officials with regards to determining the maximum number of resources required to help treat patients, and when these resources would be needed. There is a direct correlation between the total number of recorded cases and the peak number of weekly recorded cases, as well as the duration of the epidemic and the time of the peak number of cases (Figure S6 (e) and (f)).

**Figure S6:**
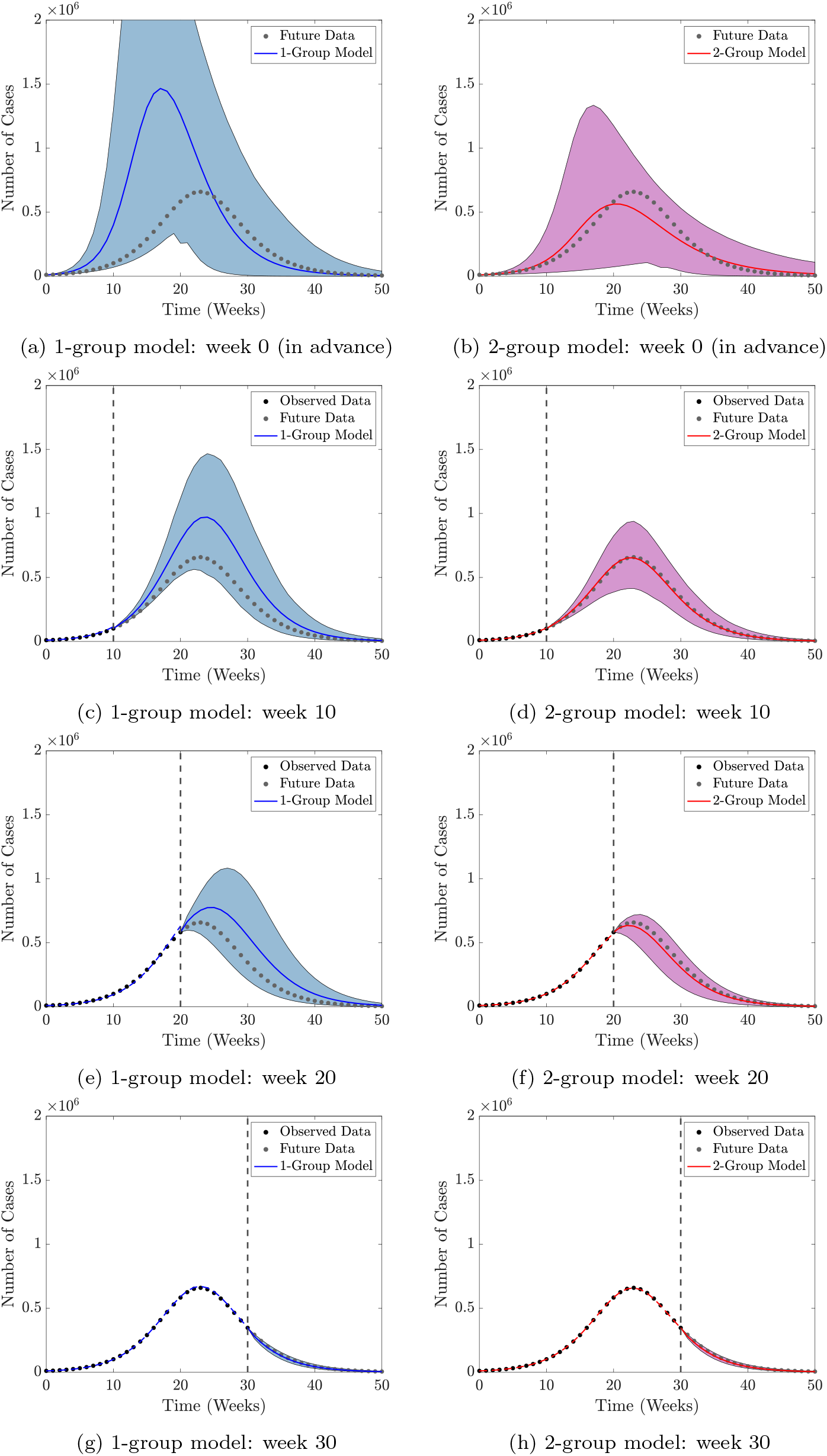
(a)-(b): The partially immune fraction of the 2-group model and the basic reproduction numbers of the 1-group and 2-group models if a second epidemic were to occur at a given time after the 2009 epidemic. (c)-(f): The total number number of (recorded) cases, the duration of the epidemic, the maximum number of weekly recorded cases, and the time at which this maximum occurs, if a second epidemic were to occur at a given time after the 2009 epidemic, as calculated by the 1-group and 2-group models. Quantities calculated using the mean parameter values estimated by fitting models to the 2009 H1N1 epidemic in Japan.

### S.9 Forecasting an Epidemic in Real-Time with Prior Information using Different Prior Parameter Distributions

We show real-time forecasts of an outbreak occurring 25 years after the 2009 epidemic (using prior information from the 2009 epidemic), if variances five times larger (Figure S7) or five times smaller (Figure S8) of both *β* and *N* are used, compared to forecasts made in Figure 6. Variances remain fixed in each figure.

**Figure S7:**
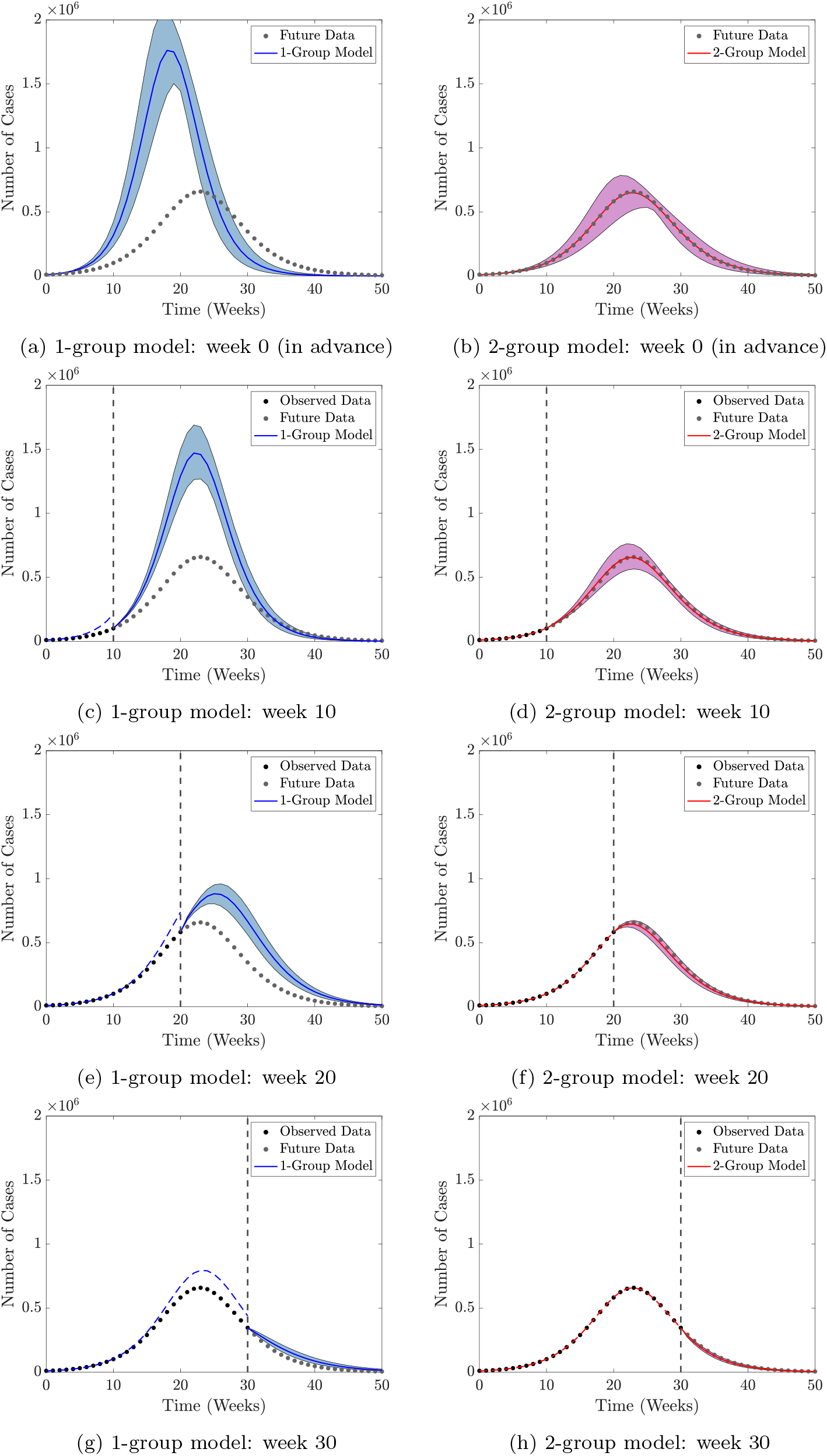
Forecasts obtained from the 1-group and 2-group models of a future epidemic occurring 25 years after the 2009 epidemic in real-time using information from previous the 2009 epidemic as in Figure 6, but with variances of the gamma distributions of *β* and *N* five times larger (details in Supplementary Information S.6).

**Figure S8:**
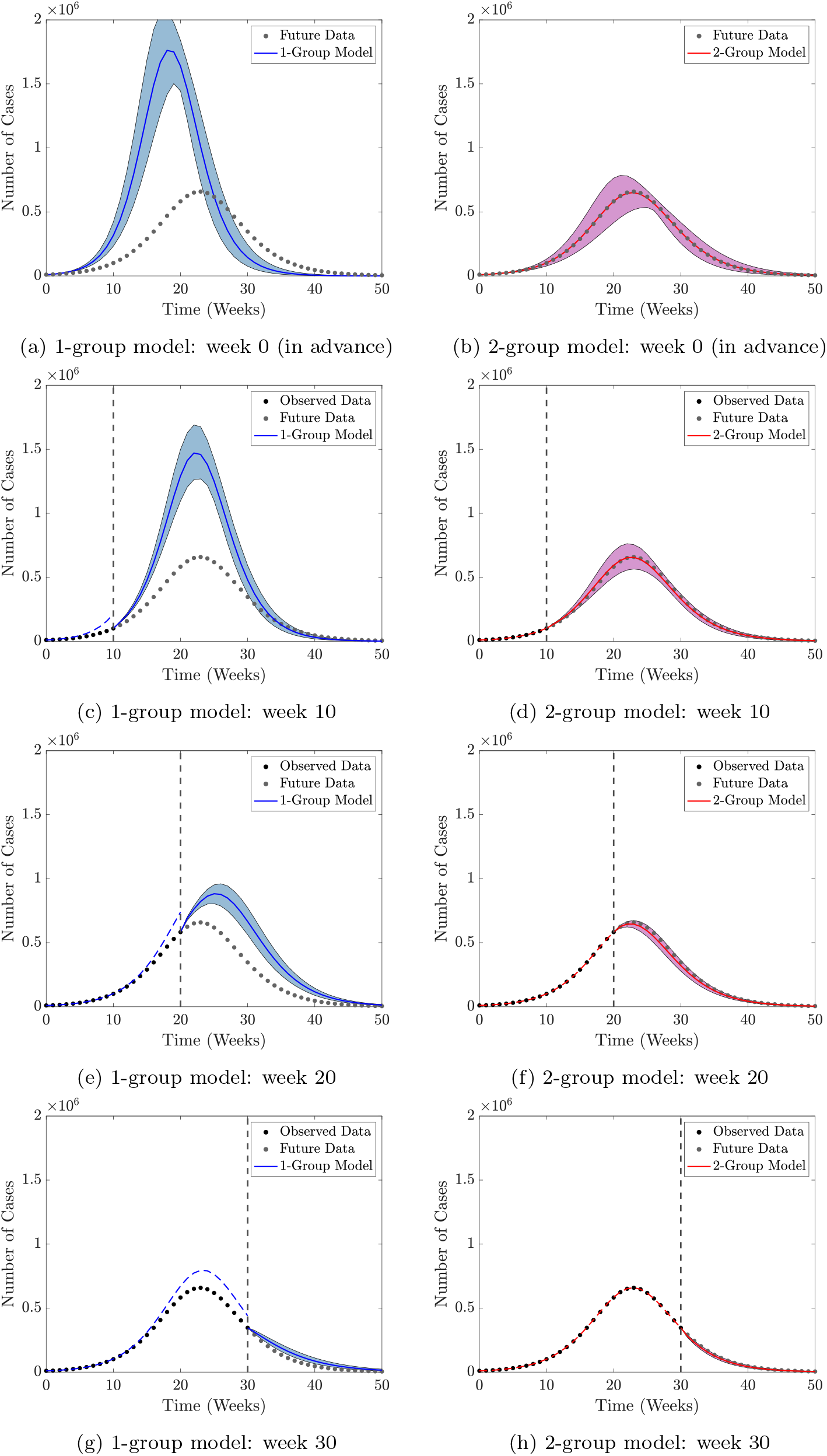
Forecasts obtained from the 1-group and 2-group models of a future epidemic occurring 25 years after the 2009 epidemic in real-time using information from previous the 2009 epidemic as in Figure 6, but with variances of the gamma distributions of *β* and *N* five times smaller (details in Supplementary Information S.6).

